# A community developed conceptual model for reducing long-term health problems in children with intellectual disability in India

**DOI:** 10.1101/2022.07.08.22277266

**Authors:** Manisha Nair, Mythili Hazarika, M Thomas Kishore, Nabarun Sengupta, Ganesh Sheregar, Hayley MacGregor, Mary Wickenden, Neel Harit Kaushik, Prarthana Saikia, Maureen Kelley, Sasha Shepperd

**Affiliations:** Nuffield Department of Population Health, University of Oxford, UK; Gauhati Medical College and Hospital, Guwahati, Assam, India; National Institute of Mental Health and Neurosciences, Bengaluru, India; Meghalaya Parents Association for Disabled, Shillong, India; National Institute for the Empowerment of Persons with Intellectual Disabilities, Secunderabad, India; Institute of Development Studies, University of Sussex, UK; Assam Don Bosco University, Sonapur, Assam, India; Indian Institute of Technology, Guwahati, Assam, India; Center for Ethics in Health Care and Department of Internal Medicine, Oregon Health & Science University; Portland, Oregon, USA

## Abstract

Children with intellectual disability (ID) have a higher risk of long-term health problems in adulthood. India has the highest prevalence of ID of any country with 1.6 million under-five children living with the condition. Despite this, compared with other children, this neglected population is excluded from mainstream disease prevention and health promotion programmes. Our objective was to develop an evidence-based conceptual framework for a needs-based inclusive intervention to reduce the risk of communicable and non-communicable diseases among children with ID in India.

From April through to July 2020 we undertook community engagement and involvement activities in ten States in India using a community-based participatory approach, guided by the bio-psycho-social model. We adapted the five steps recommended for the design and evaluation of a public participation process for the health sector.

Seventy stakeholders from ten States contributed to the project: 44 parents and 26 professionals who work with people with ID. We mapped the outputs from two rounds of stakeholder consultations with evidence from systematic reviews to develop a conceptual framework that underpins an approach to develop a cross-sectoral family-centred needs-based inclusive intervention to improve health outcomes for children with ID. A working theory of change (ToC) model delineates a pathway that reflected the priorities of the target population. We discussed the models during a third round of consultations to identify limitations, relevance of the concepts, structural and social barriers that could influence acceptability and adherence, success criteria, and integration with existing health system and service delivery.

There are currently no health promotion programmes focusing on children with ID in India despite the population being at a higher risk of developing comorbid health problems. Therefore, an urgent next step is to test the conceptual model to determine acceptance and effectiveness within the context of socio-economic challenges faced by the children and their families in the country.

## Introduction

People with intellectual disability (ID) are at an increased risk of premature ageing and death [1-3]. The adverse impact of risk factors such as malnutrition, infection, stress and anxiety, can be far reaching, leading to poor health outcomes, a lower quality of life and higher healthcare costs for people with this condition [4]. ID is “a group of etiologically diverse conditions originating during the developmental period characterised by significantly below average intellectual functioning and adaptive behaviour” [5]. Of all developmental disabilities, this is the largest contributor to years-lived-with-disability [6]. Children with ID have a 70-98% higher risk of multiple long-term health problems in adulthood [7].

India has the highest rate of ID of any country [6], a range of 1%-6.3% [8-11] is reported, compared with 0.22-1.55% globally [12]. Applying a rate of 1% [8-11] to population figures from Census 2011 [13], the total population living with ID would be at least 12 million including about 1.6 million under-five children. Prevalence is higher in some States such as the Northeast States[10]. As life expectancy of people with ID continues to increase in India [15] and in other low-and-middle income countries (LMICs) [16], preventing and reducing modifiable risk factors for long-term health problems from an early age is important to ensure healthy ageing and to prevent premature death. Evidence suggests that compared with other children, persons with ID are poor [17] and often neglected in mainstream disease prevention and health promotion programmes [2]. They are disproportionately at higher risk of developing multiple health problems due to lack of physical activity, unhealthy diet, social exclusion, poverty and the adverse effects of prolonged pharmacological treatments for co-morbidities such as epilepsy [3, 7, 18-20]. Our objective was to develop an evidence-based conceptual framework for a needs-based inclusive intervention through community consultations to reduce the risk of long-term health problems among children with ID in India.

## Methods

From April through to July 2020 we undertook community engagement and involvement (CEI) activities using a community-based participatory approach, guided by the bio-psycho-social model. This approach underpins the World Health Organisation’s (WHO’s) International Classification of Functioning Disability and Health (ICF) [21] to focus on behaviour change factors and access to healthcare services which could influence opportunities, capability and motivation of parents and children to engage with a health related intervention designed to reduce risk factors for poor health outcomes [22, 23]. Working with stakeholders we developed a conceptual framework and a working model of a ‘theory of change’ (ToC) to delineate a pathway that reflected the priorities of the target population [24]. We adapted the five steps recommended for the design and evaluation of a public participation process for the health sector [25].

### 1. Representation of a range of community stakeholders

We consulted with 70 key community stakeholders in ten States in India, eight in the northeast region that have a high prevalence of ID (Assam, Arunachal Pradesh, Manipur, Meghalaya, Nagaland, Sikkim, Mizoram, Tripura) and Delhi and Maharashtra. The stakeholders included: i) parents of children with ID; ii) professionals from health, education and social care who were engaged in caring for children and adults with ID; iii) policy makers (including commissioners), lawyers, and members of parents’ organisation and other non-governmental organisations (NGOs) working with people with ID.

### 2. A systematic approach to stakeholder interviews and consultation

We conducted three iterative rounds of one-to-one consultations with each of the 70 stakeholders. To overcome the constraints of social restrictions due to the COVID-19 pandemic, we followed guidance on e-consultations [26] to enable us to extend opportunities to as many stakeholders as possible via emails, and through tele and video-conferencing. We organised face-to-face meetings (mainly with parents) after the lockdown measures were eased in June 2020. Two psychologists (MH and NHK), parents’ representative (NS) and a PhD student (PS) from India conducted the consultations.

In the first round of consultations, we used five case studies of children with ID to initiate discussions. The case studies were compiled through an in-depth face-to-face discussion with parents prior to the COVID-19 pandemic. They provided stakeholders with a general understanding of the day-to-day living experiences, challenges, health and other concerns, fears and hopes of parents/carers of people with ID. We explored broader and more general perspectives on the:

- Day-to-day problems and needs of children with ID and their families.
- Support systems and structures available to the children and their families.
- Concerns about health and wellbeing of the children (current and future).

In round-2, we used a semi-structured topic guide to explore the stakeholders’ perception of risk factors for communicable and non-communicable diseases in children with ID and how we could start to address them within the wider context of the structural and social barriers. In round-3, we obtained critical feedback on a draft conceptual framework (described below) and a ToC model developed from the findings of round 1 and 2. In particular, the stakeholders were asked to identify limitations, assess the relevance of the concepts, acceptability, success criteria, integration with existing care pathways and health system (private and public), how these relate to existing health policies and who would be best placed to deliver the intervention.

### 3. Information generated and iteratively reviewed

Consultations undertaken in local languages were transcribed and translated to English. Transcripts of each of the 70 consultations were coded, using set and emergent themes with constant comparison [27]. The themes were assessed by study investigators (MN, SS, MH, NS, HM, MW, TKM), and ambiguous and uncertain themes were further explored with stakeholders in subsequent rounds. An iterative team coding approach was used to check interpretations and improve validity [28].

### 4. Developing a conceptual framework and theory of change

We mapped the findings from the consultations with evidence from systematic reviews to develop a conceptual framework to inform the development of a health promotion and disease prevention intervention that could reduce the known risk factors for long-term health problems in children with ID while taking into account the broader socio-economic contexts and barriers. The framework was co-developed by the study team from India and the UK working with experts from the Indian government and non-government institutions including three national bodies:

i. National Institute for the Empowerment of Persons with Intellectual Disabilities (NIEPID), Secunderabad under the Ministry of Social Justice and Empowerment, Government of India;
ii. National Institute of Mental Health and Neurosciences (NIMHANS), Bengaluru under the Ministry of Health and Family Welfare, Government of India; and
iii. PARIVAAR–National Confederation of Parents’ Organisations (NCPO) working with persons with ID.

Using our findings we worked backwards to sketch out the pathways for potential change and its preconditions, incorporating the contextual, structural and social shifts that could facilitate the intended changes within a theory of change (ToC).

### 5. Triangulation of findings for decision-making

Information from the three rounds of inter-linked and iterative discussions were triangulated to modify and refine the conceptual framework for a cross-sectoral family-centred intervention for children with ID and their parents. This was undertaken by the study team working with the same set of stakeholders who developed the initial conceptual framework (step 4 above).

## Ethics approval

CEI activities are exempt from research ethics review by the University of Oxford’s ethics committees and by the funder, National Institute for Health and Care Research (see INVOLVE statement https://www.invo.org.uk/posttypepublication/public-involvement-in-research-and-research-ethics-committee-review/). This is because such activities involve individuals in contributing to the design of a research study, but do not involve them as research participants in the study. The community consultations were nonetheless undertaken within the recommended ethical standards, including verbal consent to use quotes from stakeholders without identifying them, and their right to withdraw this approval any time without providing any reason. This work was also motivated by an ethical commitment to engaging communities in study design to ensure that the study plan reflects the priorities of the community and is responsive to the specific community context, including contextual understandings of the potential risks and benefits of research.

## Results

Despite the constraints of lockdown and social distancing regulations due to the COVID-19 pandemic in India, we achieved a good representation of parents (n=44) and other stakeholders working with children and adults with ID (n=26) from the ten States (Table-1).

**Table-1:** Characteristics of stakeholders who participated in the three rounds of consultations.

| Stakeholders and characteristics | Number |
| --- | --- |
| **Parents (total)** | **44** |
| Place of residence |  |
| Living in rural areas | 20 |
| Living in urban areas | 24 |
| Socio-economic status |  |
| Below poverty line* | 15 |
| Above poverty line | 29 |
| Education |  |
| Primary and/ or secondary education | 17 |
| Higher education/ professional | 27 |
| **Other stakeholders (total)** | **26** |
| Teachers/ educators | 5 |
| Social workers | 2 |
| Occupational and rehabilitation therapist | 1 |
| Psychologists | 8 |
| Psychiatrists | 8 |
| Policy advocates including members of civil society organisations | 2 |

\*The Planning Commission of India defines 'Below Poverty Line' households as households (average five family members) with per capita consumption expenditure of INR 672.8 on a monthly basis in rural areas and INR 859.6 in urban areas at prices prevailing in 2009-10 (Planning Commission; Government of India, 2012)

### Round-1 outputs

Three key themes emerged from the first round of consultation, the most important being the ‘future of the children’, followed by ‘inclusion’ and ‘support systems’. Almost all parents were concerned about the future care and wellbeing of their children when they would no longer be able to provide care. Marginalisation and exclusion were the factors driving insecurity and exacerbated their worries about the future of their child.

> *“I am a single parent, after my son’s birth my spouse divorced me and so we are staying with my parents. But I am suffering with insecurity when I think about his future life after my demise or in my old age. Please if it is possible you should include this issue in you[r] programme*.*” (Parent of a child with ID)*

> *“People should behave as normally as they behave with anyone else. That is the bare minimum they can do. Moreover, I expect society to be more tolerant and accommodating in general*.*” (Parent of a woman with ID)*

The majority of professionals (educators, health and social care workers) and members of parents’ organisations felt that parents should be provided with adequate knowledge and information, and be actively involved in educating and caring for the child at home. Active involvement and regular communication with healthcare providers and educators could reduce their anxiety, improve their understanding and acceptance of the child’s condition, and help to improve their mental wellbeing through support. One of the project stakeholders believed that parents are not adequately informed about their children’s condition and provided suggestions on how support can be provided at home.

> *“…they put the entire hope and aspiration on the schools and SSA [Sarva Shiksha Abhiyan] functionaries. Empowering teachers with the right skills is necessary and also empowering parents about their child’s condition and how to care at home is equally important*.*” [Special educator, SSA (Inclusive education programme)]*

Improving access to and involvement with healthcare services in both the public and private sectors were identified as important to improve the support system for children with ID and improve physical and mental health of the children and their parents. The overarching concern of parents about securing the future for the child beyond just putting food on the table and day-to-day care was an important observation.

> *“The benefit of parental and family involvement in the rehabilitation program has never been understood better than now. Professionals are beginning to realise the benefit of family approaches over merely child-centred ones. This point requires consideration*.*” (Member of a parents’ organisation)*

### Round-2 outputs

A few professionals were aware of a higher risk of communicable and non-communicable diseases in people with ID, and suggested that there was a need to improve awareness about the importance of early interventions to reduce future risk of diabetes, cardiovascular diseases, mental health problems, tuberculosis, COVID-19 and other infections. An increased risk of physical health problems was attributed to a range of factors that included malnutrition, infection, poverty, poor access to healthcare services, societal factors such as those related to stigma and low inclusion, behavioural and lifestyle factors, and a lack of awareness and knowledge about health promotion and disease prevention among parents and individuals with ID.

> *“Children and persons with ID besides being faced with known health concerns, are more vulnerable and susceptible to other illnesses due to lower immunity levels and also because they are not aware and equipped with required knowledge to take good care of their general health – both mental and physical*.*” (Deputy Director, Civil Society organisation)*

> *“Another critical concern is the lack of acknowledgment among parents and other caregivers about the sexual needs, identities and expressions of children and adolescents with ID. Championing for the same could be viewed with a lens of censorship, when societal norms dictate that these are ‘private’ matters. As a result, these are not addressed timely and adequately, thereby impacting the mental well-being of people with ID and sometimes also resulting in unwanted changes in their behaviours like resorting to violence and so on*.*” (Deputy Director, Civil Society organisation)*

Parents of children with ID mostly reported not being aware of any increased risk of long-term health problems, but mentioned that they would do everything required to prevent it. The statements quoted above also point to a lack of awareness of inclusive approaches amongst parents, professionals and the wider society. An educator raised concern about the lack of health promotion and disease prevention programmes for children with ID.

> *“Government has taken up education of these children as a priority, whereas health [promotion] still remains to be addressed. Maybe it will be prioritised in the next phase. NGOs are also focusing more on education than health. I would press for a health related programme*.*” (Educator, inclusive education programme)*

The common recommendations for reducing the increased risk of long-term health problems were to increase physical activity levels in children with ID by integrating this with daily activities and to improve their nutrition. Social workers supporting children with ID suggested that the intervention should be tailored to the needs of children from different socio-economic backgrounds, and take into account the severity of disability and any co-existing conditions. We were advised to keep the intervention simple, with separate components for undernutrition and obesity, to ensure it is sustainable and build on the existing Early Intervention Services for children 0-3 years offered by the Rashtriya Bal Swasthya Karyakram of the Ministry of Health and Family Welfare, Government of India [29]. This will also facilitate an inclusive approach, as the intervention will be embedded in a mainstream programme for all children. Experts from the three national bodies advised us to target the age group of 4-10 years to support continuity of care and integration with the existing policies and programmes.

> *“Early childhood interventions are necessary and should be continued as part of the programme. If a child does not know how to brush, how can you improve dental hygiene?” (President, National parents’ organisation)*

### Conceptual framework that underpins an intervention to reduce risk factors for long-term health problems

A report on the state-level burden of disease in the general population highlights the triple-burden of infectious and lifestyle related diseases, and mental health problems among the adult population in India. These are associated with risk factors such as malnutrition, high blood pressure, high blood sugar, stress and anxiety, which are in turn related to unhealthy diet, tobacco, alcohol and drug use, socio-economic factors, and unsafe drinking water, sanitation, and hygiene [30]. A review of systematic reviews indicated that interventions to increase physical activity had a large beneficial effect on physical and psychosocial health moderated by age and level of disability [31] although this evidence mainly came from high-income countries. Sustained behaviour change was higher when combined with a healthy lifestyle and supportive structural and socio-economic environments [32]. Community-based programmes that provided meaningful parent participation (e.g. through having fun, role models) reduced the burden of caregiving [33]. Integrated mental health counselling services within a health promotion programme for children with ID was found to be effective in reducing stress and anxiety among parents [34]. Evidence also supported collaboration with adults with ID in all aspects of research to improve the implementation and sustainability of programmes [35].

Combining the evidence with the findings from the stakeholder consultations, we developed a conceptual framework that covered an enabling environment, structural and socio-economic determinants, integration with health and social care, intermediate modifiable risk factors and the long-term burden of multiple health problems in later life (Figure-1). This led to the identification of five evidence-based core components to underpin a family-centred, cross-sectoral, community-based intervention for children with ID aged 4 to 10 years:

**Figure-1:**
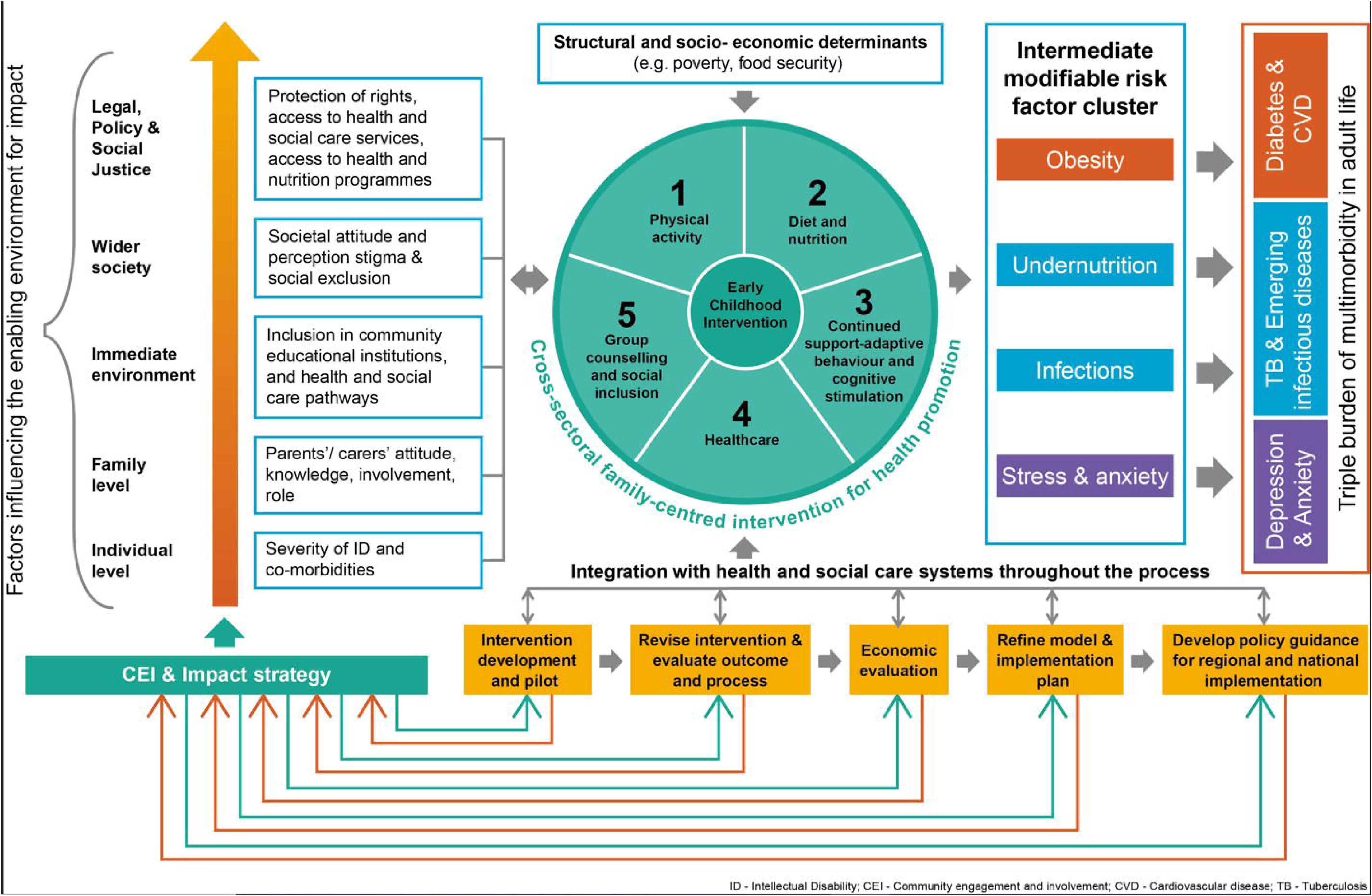
Conceptual framework for a community-based intervention for preventing and reducing risk of long-term health problems in children with intellectual disability

i. A physical activity programme for the children.
ii. A guide to a healthy diet to provide good nutrition to the children using locally available and affordable food.
iii. Training parents to continue regular adaptive behavioural and cognitive stimulation, including in everyday activities alongside other children who do not require specialist resources.
iv. Education and training to improve parent’s knowledge of accessing healthcare services.
v. Group counselling for parents and children to increase their capability to address behavioural issues, individual barriers to exercise and how to follow a healthy diet, improve emotional and mental wellbeing, and to encourage social inclusion.

We developed a ToC model (Figure-2) to identify what a change in risk for longer-term health problems would mean for our target population of children and their parents, and the necessary preconditions for this. This provided guidance for further refinement of the concepts along with the stakeholders before testing their effect on the modifiable risk factors. The cross-sectoral family-centred programme is not a standalone intervention, but will need to be supported by an inclusive enabling environment (Figure-1). Therefore, it will be essential to identify and mitigate social and health system barriers, and actively promote community mobilisation to integrate the intervention within existing health and social care services (Figure-2). While the intervention could achieve the immediate outcomes within a short period (example improving physical activity, diet, and access to healthcare, and reducing stress and anxiety), it will take longer to observe an impact on reducing the long-term health problems in children with ID. This is clearly acknowledged by the accountability ceiling and the plan to transfer ownership of the intervention to the three national apex bodies engaged in developing the conceptual models. The ToC model will evolve as the intervention is tested and implemented, along with growing partnerships and changes in political will.

**Figure-2:**
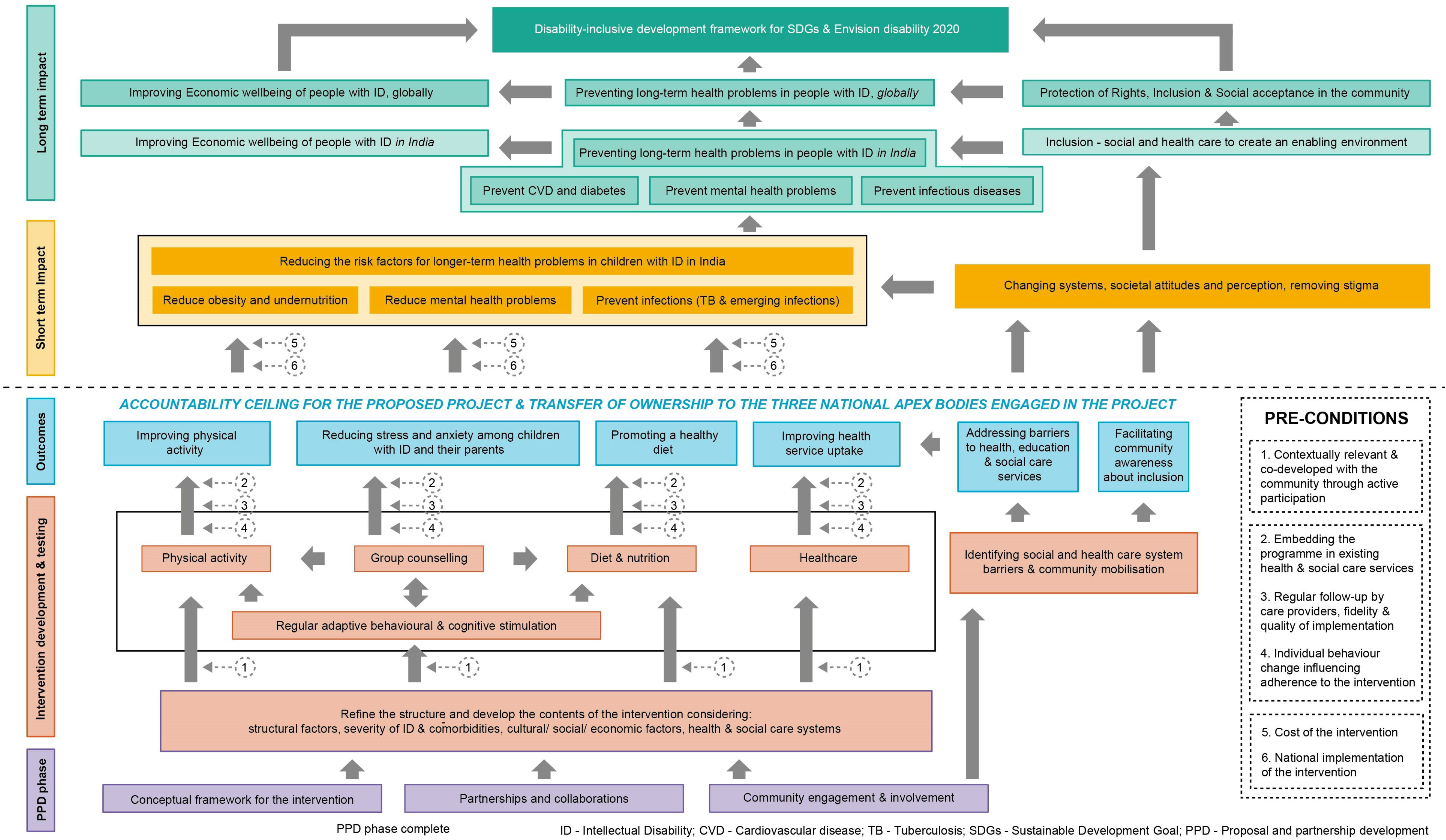
Theory of change (ToC) model

### Round-3 outputs

Stakeholders voiced strong support for the conceptual framework that inform the intervention to guide the planning and delivery of services for this population, and agreed that *“it will mark a beginning” (Educator, Inclusive education programme)* and promote the *“value of life of the child” (Carer of a child with ID)*.

> *“The programme would be a good start of a holistic intervention. Done with perseverance, it can positively impact the child’s coping skills, health and well-being and relieve stress in the family as the child begins to benefit from inputs received*.*” (Deputy Director, Civil Society organisation)*

Importantly, it was suggested that we develop a contextually relevant intervention and an enabling environment that will address the challenges and potential barriers to adherence to the intervention components.

While several stakeholders suggested that grass-roots level public sector health and nutritional workers such as Anganwadi workers (village nutrition worker) and community health workers (Accredited Social Health Activist (ASHA)) should be trained to deliver the intervention, others were worried about the negative impact on quality and sustainability if it is delivered by this already over-burdened workforce. It was felt that care providers employed by government and non-government institutions, such as the health system, nutrition and social welfare sectors and schools for children with disabilities would be better placed to deliver the intervention due to their understanding of the children’s complex needs. The district disability rehabilitation centres [36] could coordinate outreach, delivery of the intervention and follow-up. The trained workers in these centres are already working towards improving societal awareness and attitudes, community inclusion of people with disability, and school enrolment, and helping families access services, benefits, and support for their children [37].

## Discussion

Through the extensive community engagement and involvement (CEI) activities we developed a conceptual model for a community-based, cross-sectoral, family-centred inclusive intervention to prevent long-term health problems in children with ID by targeting modifiable risk factors from an early age (4-10 years), and address the stigma and social exclusion that exacerbate the course and experience of health problems. The intervention has the potential to address some of the modifiable risk factors by promoting a healthy lifestyle, preventing under and over nutrition, and improving healthcare access for children, and reducing stress and anxiety among the children and their parents in the short term. Although stakeholders did not mention the inclusion of children with disability into the mainstream health promotion activities, we added this to our conceptual model and ToC to align with the sustainable and inclusive approach recommended by UNCRPD [38, 39].

Health promotion for children with ID can be implemented through the inclusive education programme. However, this will require attention to the structural, social and health system barriers, especially those related to stigma that limit inclusion. The conceptual framework aligns with the recommendation on health and early intervention for children with ID included in the India’s National Policy for Persons with Disabilities (No.3-1/1993-DD.III, 2006), which promotes an inclusive approach. Our ToC model outlines the pathways through which the intervention together with efforts to creating an enabling environment could translate into reducing communicable and non-communicable diseases, and mental health problems, thereby preventing morbidity and mortality in people with ID in India in the longer term. The framework can be adapted to other settings.

Wider activism for societal change, recognition of disability rights to an equal and inclusive world and economic support through social grants, etc will be required to address the socio-economic barriers to health. CEI being at the core of the work ensured acceptance and strong support from the community, from parents/carers of children with ID, adults with ID, educators, policy makers, clinicians, and civil society organisations.

People with ID are also socio-economically the most vulnerable population in India [12] and other LMICs [17] due to the limited opportunities for employment and income. Many have co-existing conditions (such as epilepsy) that require life-long treatment associated with extra expense for families. It is estimated that disability increases the cost of living by more than 30-40% of average income [40]. Thus, developing further morbidities (communicable and non-communicable diseases) would have far worse consequences for people with ID, than for others. Considering that healthcare spending in India is mostly out-of-pocket [41] and people with ID often have additional costs related to transportation and needing carers to accompany them [2], developing long-term health problems would add to the burden of ongoing costs resulting in further impoverishment of this vulnerable population. Therefore, even a small effect of the intervention will have a large positive impact on individuals, families and the society in the long term. However, the conceptual model needs to be tested and further refined in order to develop an effective and sustainable intervention together with an enabling inclusive environment that can be scaled-up across India and also adapted by other LMICs. Although the alignment of the conceptual framework with India’s National Policy for Persons with Disabilities will support integration [42], further inclusion within a wide range of existing government programmes would be needed to facilitate implementation, ownership and sustainability [43].

The conceptual model targets behaviour change, using the bio-psycho-social model and WHO’s ICF [44], to support sustainable uptake. This will be further supported by the early provision of the intervention, plus an element of standardisation of simple to deliver core elements that maintain flexibility according to local circumstances. Research that tests cost-effectiveness and implementation by parents and educators is required [45]. Another important point in changing behaviour (in our case physical activity, diet, accessing healthcare) is to set expectations that are ‘attainable’ and ‘reasonable’[45] within an enabling environment.

## Strengths and limitations

A community-based participatory approach was used for consulting and actively engaging stakeholders in co-developing a ‘locally conceptualised’ needs-based intervention to achieve meaningful results that maximises the benefits for children with ID and their parents. Our structured approach could be used to undertake other CEI activities in India and other LMICs, where communities and individual members of the public are not commonly engaged in developing an intervention. Consultations are thought to be more effective if they are conducted in open fora where the public can debate and challenge emerging concepts. Although we could not reach out to a wider community because of the social restrictions related to the pandemic, we minimised the chances of developing a biased model by listening to and evaluating multiple perspectives along with experts from the three national bodies who have several years of experience working with children and adults with ID in the Indian context. Another potential limitation is that systematic reviews used to inform the evidence base of the conceptual framework was from mainly high-income countries. Nevertheless, the evidence was a necessary starting point. Evidence generated from a LMIC context is required to strengthen the conceptual model, guide implementation and assess how best to align with the “disability-inclusive development” framework [46] when substantial socio-economic and health system challenges exist. Our work reinforces the United Nation’s Convention on the Rights of Persons with Disabilities (UNCRPD) and the Sustainable Development Goals’ (SDGs) commitment to “leave no one behind”. It also contributes towards building inclusive societies and institutions and to the goals of ‘Envision disability 2020’ [47].

## Conclusion

There is an urgent need for appropriate strategies to reduce long-term health problems in children with ID as they move into earlier and later adulthood. Interventions guided by the conceptual framework have the potential to reduce and prevent long-term health problems in millions of children living with ID in India, and potentially a higher number globally. Considering that there are no health promotion programmes, which proactively include children with ID in LMICs, an urgent next step is to test the conceptual model together with actions for improving the enabling environment in order to determine its acceptance and effectiveness.

## Data Availability

Our qualitative data includes transcripts from a public engagement and involvement work. Even if we remove the names and other personal details, the data would be still potentially identifiable of the persons/organisations including government representatives whose views might be considered professionally and politically sensitive. This is particularly true for our work that focuses on intellectual disability and includes quotes related to stigma and social injustice. So, to protect confidentiality we did not seek consent from the participants to share data in an open access repository. Participants consented to including quotes in scientific reports, publications and presentations, but not to have entire transcripts made publicly available or available to a third party. We would be happy to consider requests for data from interested individuals or organisations who approach the corresponding author provided we could work out ways to share non-identifiable information.

## Acknowledgement

We thank all stakeholders who participated in the CEI activities and shared their views, opinions, ideas, and reviewed the conceptual framework and the ToC model. We also thank Professor Suresh Bada Math, Mr Arman Ali, Dr Shyamanta Das, Dr Mara Violato, Dr Binukumar Bhaskarapillai, Dr Louise Linsell and Professor John Vijay Sagar Kommu who were co-investigators for the wider project. They did not specifically contribute to the work included in this paper, but provided broader perspectives for developing the overall project.

## Author contributions

MN co-developed the concept for the CEI work, analysed the information, conducted a review of systematic reviews, co-developed the methodology, co-developed the conceptual framework and the ToC model, and wrote the first draft of the paper. MH co-developed the concept for the CEI work, facilitated stakeholder interviews, contributed to analysing the information, co-developed the conceptual framework and the ToC model. MTK co-developed the concept for the CEI work, co-developed the methodology, contributed to analysing the information, co-developed the conceptual framework and the ToC model, and contributed to writing the paper. NS co-developed the concept for the CEI work, conducted stakeholder interviews, co-developed the conceptual framework and the ToC model, and edited the paper. GS co-developed the concept for the CEI work, contributed to analysing the information, co-developed the conceptual framework and the ToC model. HM and MW co-developed the concept for the CEI work, co-developed the methodology, co-developed the conceptual framework and the ToC model and edited the paper. NHK conducted stakeholder interviews, contributed to analysing the information, and coordinated the CEI activities. PS conducted stakeholder interviews and contributed to analysing the information. MK provided ethical expertise to the project, contributed to analysing the information and edited the paper. SS co-developed the concept for the CEI work, co-developed the methodology, contributed to analysing the information, co-developed the conceptual framework and the ToC model, and edited the paper.

## Funding

The CEI work included in this paper was funded by a National Institute for Health and Care Research (NIHR), RIGHT Call 3 Proposal and Partnership Development Award (Ref: NIHR201720). MN is funded by a Medical Research Council Transition Support Award (Ref: MR/W029294/1). The funders had no role in the study design, data collection, analysis, or writing the paper.

## Competing interests statement

The authors declare that they have no competing interests.

## Notes

### Competing Interest Statement

The authors have declared no competing interest.

## References

1. Ali A, Hassiotis A, Strydom A, King M. Self stigma in people with intellectual disabilities and courtesy stigma in family carers: a systematic review. Research in developmental disabilities. 2012;33(6):2122–40. Epub 2012/07/13. doi: 10.1016/j.ridd.2012.06.013. PubMed PMID: 22784823.

2. World Health Organisation. Disability and health: WHO; 2018 [cited 2020 03 August]. Available from: https://www.who.int/news-room/fact-sheets/detail/disability-and-health.

3. Tyrer F, Dunkley AJ, Singh J, Kristunas C, Khunti K, Bhaumik S, et al. Multimorbidity and lifestyle factors among adults with intellectual disabilities: a cross-sectional analysis of a UK cohort. Journal of intellectual disability research : JIDR. 2019;63(3):255–65. Epub 2018/11/30. doi: 10.1111/jir.12571. PubMed PMID: 30485584.

4. Sciences AoM. Multimorbidity: a priority for global health research. London: Academy of Medical Sciences, 2018.

5. World Health Organisation. ICD-11 for Mortality and Morbidity Statistics (Version : 04 / 2019) Geneva: WHO; [cited 2019 16-December].

6. Olusanya BO, Davis AC, Wertlieb D, Boo N-Y, Nair MKC, Halpern R, et al. Developmental disabilities among children younger than 5 years in 195 countries and territories, 1990–2016: a systematic analysis for the Global Burden of Disease Study 2016. The Lancet Global Health. 2018;6(10):e1100–e21. doi: 10.1016/S2214-109X(18)30309-7.

7. Kinnear D, Morrison J, Allan L, Henderson A, Smiley E, Cooper SA. Prevalence of physical conditions and multimorbidity in a cohort of adults with intellectual disabilities with and without Down syndrome: cross-sectional study. BMJ open. 2018;8(2):e018292. Epub 2018/02/13. doi: 10.1136/bmjopen-2017-018292. PubMed PMID: 29431619; PubMed Central PMCID: PMCPMC5829598.

8. Kishore M T, Udipi GA, Seshadri SP. Clinical practice guidelines for assessment and management of intellectual disability. Indian J Psychiatry. 2019;61(Suppl S2):194–210.

9. Chavan BS, Rozatkar AR. Intellectual disability in India: Charity to right based. Indian journal of psychiatry. 2014;56(2):113–6. doi: 10.4103/0019-5545.130477. PubMed PMID: 24891695.

10. Sagar R, Dandona R, Gururaj G, Dhaliwal RS, Singh A, Ferrari A, et al. The burden of mental disorders across the states of India: the Global Burden of Disease Study 1990–2017. The Lancet Psychiatry. doi: 10.1016/S2215-0366(19)30475-4.

11. Gururaj G, Varghese M, Benegal V, Rao GN, Pathak K, Singh LK, et al. National Mental Health Survey of India, 2015-16: Summary. Bengaluru: National Institute of Mental Health and Neuro Sciences, NIMHANS, 2016 Contract No.: Publication No. 128.

12. National Institute for the Empowerment of Persons with Intellectual Disabilities (Divyangjan). 35th Annual Report 2018-19. Secunderabad, Telangana, India: Department of Empowerment of Persons with Disabilities (Divyangjan), Ministry of Social Justice and Empowerment, Government of India, 2019.

13. Office of the Registrar General & Census Commissioner India. 2011 Census Data New Delhi: Ministry of Home Affairs, Government of India; [cited 2020 08 September].

14. Lakhan R, Ekúndayò OT, Sharma M. Epilepsy, Behavioral Problems, and Intellectual Disability among Children in India: Conundrums and Challenges. J Neurosci Rural Pract. 2018;9(1):1–2. doi: 10.4103/jnrp.jnrp_477_17. PubMed PMID: 29456334.

15. Menon DK, Kishore MT, Sivakumar T, Maulik PK, Kumar D, Lakhan R, et al. The National Trust: A viable model of care for adults with intellectual disabilities in India. Journal of intellectual disabilities : JOID. 2017;21(3):259–69. Epub 2017/08/17. doi: 10.1177/1744629517709832. PubMed PMID: 28812964.

16. Evenhuis H, Henderson CM, Beange H, Lennox N, Chicoine B. Healthy Ageing - Adults with Intellectual Disabilities: Physical Health Issues. Geneva, Switzerland: World Health Organization, 2000.

17. World bank. Disability inclusion [updated 15 May 2020; cited 2020 01 September]. Available from: https://www.worldbank.org/en/topic/disability#:∼:text=Results-,One%20billion%20people%2C%20or%2015%25%20of%20the%20world’s%20population%2C,million%20people%2C%20experience%20significant%20disabilities.

18. Robertson J, Hatton C, Emerson E, Baines S. Prevalence of epilepsy among people with intellectual disabilities: A systematic review. Seizure. 2015;29:46–62. doi: https://doi.org/10.1016/j.seizure.2015.03.016.

19. van Timmeren EA, van der Schans CP, van der Putten AA, Krijnen WP, Steenbergen HA, van Schrojenstein Lantman-de Valk HM, et al. Physical health issues in adults with severe or profound intellectual and motor disabilities: a systematic review of cross-sectional studies. Journal of intellectual disability research : JIDR. 2017;61(1):30–49. Epub 2016/05/28. doi: 10.1111/jir.12296. PubMed PMID: 27228900.

20. Mitter N, Ali A, Scior K. Stigma experienced by families of individuals with intellectual disabilities and autism: A systematic review. Research in developmental disabilities. 2019;89:10–21. Epub 2019/03/16. doi: 10.1016/j.ridd.2019.03.001. PubMed PMID: 30875608.

21. World Health Organisation. International Classification of Functioning, Disability and Health (ICF) Geneva: WHO; [cited 2019 16-December]. Available from: https://www.who.int/classifications/icf/en/.

22. Higginbottom G, Liamputtong P. What is participatory research? why do it? In: Higginbottom G, Liamputtong P, editors. Participatory Qualitative Research Methodologies in Health. 55 City Road, London: SAGE Publications Ltd; 2015.

23. Hebert JR, Brandt HM, Armstead CA, Adams SA, Steck SE. Interdisciplinary, translational, and community-based participatory research: finding a common language to improve cancer research. Cancer epidemiology, biomarkers & prevention : a publication of the American Association for Cancer Research, cosponsored by the American Society of Preventive Oncology. 2009;18(4):1213–7. Epub 2009/04/02. doi: 10.1158/1055-9965.Epi-08-1166. PubMed PMID: 19336548; PubMed Central PMCID: PMCPMC2679168.

24. De Silva MJ, Breuer E, Lee L, Asher L, Chowdhary N, Lund C, et al. Theory of Change: a theory-driven approach to enhance the Medical Research Council’s framework for complex interventions. Trials. 2014;15(1):267. doi: 10.1186/1745-6215-15-267.

25. Abelson J, Forest P-G, Eyles J, Smith P, Martin E, Gauvin F-P. Deliberations about deliberative methods: issues in the design and evaluation of public participation processes. Social Science & Medicine. 2003;57(2):239–51. doi: https://doi.org/10.1016/S0277-9536(02)00343-X.

26. Medaglia R. eParticipation research: Moving characterization forward (2006–2011). Government Information Quarterly. 2012;29(3):346–60. doi: https://doi.org/10.1016/j.giq.2012.02.010.

27. Braun V, Clarke V. Using thematic analysis in psychology. Qualitative Research in Psychology. 2006;3(2):77–101. doi: 10.1191/1478088706qp063oa.

28. Morse JM, Barrett M, Mayan M, Olson K, Spiers J. Verification strategies for establishing reliability and validity in qualitative research. International journal of qualitative methods. 2002;1(2):13–22.

29. Ministry of Health & Family Welfare GoI. Rashtriya Bal Swasthya Karyakram (RBSK): Ministry of Health & Family Welfare, Government of India; [cited 2021 10 June]. Available from: https://rbsk.gov.in/RBSKLive/.

30. Indian Council of Medical Research, Public Health Foundation of India, Institute for Health Metrics and Evaluation. India: Health of the Nation’s States - The India State-Level Disease Burden Initiative. New Delhi, India: ICMR, PHFI, IHME, 2017.

31. Kapsal NJ, Dicke T, Morin AJS, Vasconcellos D, Maiano C, Lee J, et al. Effects of Physical Activity on the Physical and Psychosocial Health of Youth With Intellectual Disabilities: A Systematic Review and Meta-Analysis. Journal of physical activity & health. 2019:1–9. Epub 2019/10/06. doi: 10.1123/jpah.2018-0675. PubMed PMID: 31586434.

32. Hassan NM, Landorf KB, Shields N, Munteanu SE. Effectiveness of interventions to increase physical activity in individuals with intellectual disabilities: a systematic review of randomised controlled trials. Journal of intellectual disability research : JIDR. 2019;63(2):168–91. Epub 2018/11/09. doi: 10.1111/jir.12562. PubMed PMID: 30407677.

33. Willis C, Girdler S, Thompson M, Rosenberg M, Reid S, Elliott C. Elements contributing to meaningful participation for children and youth with disabilities: a scoping review. Disability and rehabilitation. 2017;39(17):1771–84. Epub 2016/07/22. doi: 10.1080/09638288.2016.1207716. PubMed PMID: 27442686.

34. Hinckson EA, Dickinson A, Water T, Sands M, Penman L. Physical activity, dietary habits and overall health in overweight and obese children and youth with intellectual disability or autism. Research in developmental disabilities. 2013;34(4):1170–8. doi: https://doi.org/10.1016/j.ridd.2012.12.006.

35. Di Lorito C, Bosco A, Birt L, Hassiotis A. Co-research with adults with intellectual disability: A systematic review. Journal of applied research in intellectual disabilities : JARID. 2018;31(5):669–86. Epub 2017/12/13. doi: 10.1111/jar.12435. PubMed PMID: 29231285.

36. World Health Organisation. Disability: Community-based rehabilitation (CBR) Geneva: WHO; [cited 2020 13 September]. Available from: https://www.who.int/disabilities/cbr/en/#:∼:text=Community%2Dbased%20rehabilitation%20(CBR)%20was%20initiated%20by%20WHO%20following,ensure%20their%20inclusion%20and%20participation.

37. Girimaji SC. Intellectual disability in India: the evolving patterns of care. International Psychiatry. 2011;8(2).

38. Hashemi G, Kuper H, Wickenden M. SDGs, Inclusive Health and the path to Universal Health Coverage. Disability and The Global South. Disability and The Global South. 2017;4(1):1088–111.

39. UN TU-. United Nations convention on the rights of persons with disabilities 2007 [cited 2022 08 June]. Available from: https://www.un.org/development/desa/disabilities/convention-on-the-rights-of-persons-with-disabilities.html.

40. United Nations Department of Economic and Social Affairs. Disability and Development Report: Realizing the Sustainable Development Goals by, for and with persons with disabilities. New York: United Nations, 2018.

41. Pandey A, Ploubidis GB, Clarke L, Dandona L. Trends in catastrophic health expenditure in India: 1993 to 2014. Bull World Health Organ. 2018;96(1):18–28. Epub 2017/11/30. doi: 10.2471/BLT.17.191759. PubMed PMID: 29403097.

42. Government of India. National Policy for Persons with Disabilities (No.3-1/1993-DD.III). New Delhi: Ministry of Social Justice and Empowerment

43. NIMH and IIPA. Disability management in India: challenges and commitments. Mohapatra CS, editor. Secunderabad: National Institute for the Mentally Handicapped; 2004.

44. Kostanjsek N. Use of The International Classification of Functioning, Disability and Health (ICF) as a conceptual framework and common language for disability statistics and health information systems. BMC Public Health. 2011;11 Suppl 4(Suppl 4):S3–S. doi: 10.1186/1471-2458-11-S4-S3. PubMed PMID: 21624189.

45. Zucker GH. Intervention Strategies for Pre-School Students with Special Needs. Forum on Public Policy, 2010.

46. UNDP. Disability Inclusive Development in UNDP: Guidance and entry points. New York: United Nations Development Programme (UNDP), Bureau for Policy and Programme Support, 2018. 47.

47. United Nations Department of Economic and Social Affairs - Disability. #Envision2030: 17 goals to transform the world for persons with disabilities [cited 2020 22 June]. Available from: https://www.un.org/development/desa/disabilities/envision2030.html.

